# Prevalence and Risk Factors for Acute Posttraumatic Stress Disorder during the COVID-19 Outbreak

**DOI:** 10.1101/2020.03.06.20032425

**Authors:** Luna Sun, Zhuoer Sun, Lili Wu, Zhenwen Zhu, Fan Zhang, Zhilei Shang, Yanpu Jia, Jingwen Gu, Yaoguang Zhou, Yan Wang, Nianqi Liu, Weizhi Liu

**Author notes:** **Corresponding author:** Weizhi Liu, PhD; MD, Lab for Post-traumatic Stress Disorder, Faculty of Psychology and Mental Health, Naval Medical University, 800 Xiangyin Road, Shanghai 200433, China. these authors contributed equally to this work (Luna Sun, Zhuoer Sun, Lili Wu, Zhenwen Zhu).

## Abstract

**Background:** To exam the prevalence of and risk factors for acute posttraumatic stress disorder (PTSD) in Chinese people shortly after the COVID-19 outbreak.

**Methods:** An online questionnaire survey was conducted between 30 January and 3 February, 2020. The survey included two self-administered questionnaires: one was designed to require participants’ personal information (gender, age, education background), current location, recent epidemic area contact history, the classification of population, and subjective sleep quality; the other was the PTSD Checklist for DSM-5 (PCL-5).

**Results:** A total of 2091 Chinese participated in this study. The prevalence of PTSD among the Chinese public one month after the COVID-19 outbreak was 4.6%. Multiple linear regression analysis revealed that gender (*p* < 0.001), epidemic area contact history (*p* = 0.047), classification of population (*p* < 0.001), and subjective sleep quality (*p* < 0.001) could be regarded as predictor factors for PTSD.

**Limitations:** First, the majority of participants in this study were the general public, and confirmed or suspected patients being a small part. Second, the measurement of PTSD might be vulnerable to selection bias because of an online self-report study, such as participants’ recruitment. Third, the prevalence of PTSD in this study was estimated by an online questionnaire rather than a clinical interview.

**Conclusions:** The results revealed that some Chinese showed acute PTSD during the COVID-19 outbreak. Therefore, comprehensive psychological intervention needs further implementation. Furthermore, females, people who having recent epidemic area contact history, those at high risk of infection or with poor sleep quality deserve special attention.

## Introduction

A novel coronavirus, now named as severe acute respiratory syndrome coronavirus 2 (SARS-CoV-2) by the Coronavirus Study Group (CSG) (Alexander et al., 2020), was first reported in China in December 2019 (The 2019-nCoV Outbreak Joint Field Epidemiology Investigation Team and Li, 2020; Zhu et al., 2020). On 11 February, 2020, the disease caused by the novel coronavirus was officially named as coronavirus disease 2019 (COVID-19) by World Health Organization (WHO)(WHO, 2020). The outbreak of COVID-19 was unique in its rapidity of transmission, which has become a global health emergency within just a few weeks (Wang et al., 2020). Moreover, the number of confirmed and suspected cases has rapidly risen not only in China but also in other countries (Munster et al., 2020). According to reports from WHO, there have been 93,194,922 confirmed cases globally by 17 January, 2021, including 2,014,729 deaths (WHO, 2021). As the outbreak continue to escalate, it has raised both public health concerns and tremendous psychological distress, especially the development of posttraumatic stress disorder (PTSD), which will lead to chronic symptoms, such as intrusive memories, avoidance behaviors, irritability, and emotional numbing.

PTSD is a stress-related psychological problem that occurs in someone who has experienced or witnessed a life-threatening traumatic event, which has contributed to a substantial burden on individuals and the society (Kessler, 2000). According to previous literature, exposure to traumatic events is the immediate cause of PTSD and is essential in diagnosing related symptoms (Lin et al., 2007; Wu et al., 2005). Based on Life Events Checklist for DSM-5 (LEC-5) (Weathers et al., 2013a), being involved in a life-threatening illness might be a traumatic experience. Previous research on the mental health of medical staff involved in the 2003 severe acute respiratory syndrome (SARS) outbreak found that about 10% of the sample reported high levels of PTSD symptoms (Wu et al., 2009). Another study showed that about 2% Chinese university students met symptom criteria for PTSD during the 2009 H1N1 (Influenza A virus subtype H1N1) pandemic (Xu et al., 2011). Research on the long-term psychiatric morbidities among SARS survivors revealed that PTSD was the most common long-term psychiatric disorder, and the cumulative incidence of PTSD in the 30 months since the SARS outbreak was 47.8% (Mak et al., 2009). Furthermore, it has been demonstrated that COVID-19 can be identified as a traumatic event and can cause PTSD symptoms in the general public (Forte et al., 2020). In conclusion, early detection and intervention for PTSD is necessary in the COVID-19 pandemic as well as future major life-threatening disease outbreaks.

The present study was to explore two research questions regarding acute PTSD symptoms among Chinese one month after the COVID-19 outbreak. Firstly, the severity and prevalence of PTSD among Chinese people during the outbreak were estimated. Secondly, related risk factors for PTSD were also investigated in this study. Existing literature pointed out that gender, age, education background, the degree of exposure, having been infected, and having family, friends or acquaintances infected could be identified as predictors of PTSD during outbreaks of infectious diseases, such as SARS and H1N1 (Lee et al., 2006; Xu et al., 2011). Additionally, sleep disturbances are seriously prominent in trauma-related diseases (Spoormaker and Montgomery, 2008). Therefore, gender, age, education background, current location, recent epidemic area contact history, risk of infection (based on classification of population), and subjective sleep quality were hypothesized as associated factors of PTSD in this study. Besides, since new confirmed and suspected cases rapidly grew without any hint of decline in China from the end of January 2020 to the beginning of February 2020, people were bombarded with mass information about the pandemic, which might affect their psychological condition drastically. The date participants filling out the questionnaire (identified as reporting date) was also considered in the study. The findings of this study will offer useful psychological guidance to people who are in need, and provide helpful insight into planning for future outbreaks of emerging infectious diseases.

## Methods

### Participants

This study was conducted in the mainland of China (including 31 provinces, cities and autonomous regions, and 2 special administrative regions) by using self-administered questionnaires between 30 January and 3 February, 2020. All participants were recruited online, which has been shown benefits in feasibility and availability (Batterham and J., 2014; Juraschek et al., 2018; Leach et al., 2017; Quach et al., 2013). The inclusion criteria were: 1) living in the mainland of China during the COVID-19 outbreak; 2) no history of mental illness; 3) no cognitive impairment. It took approximately 15 minutes to complete the questionnaires. Participants who responded less than 2 minutes or more than 30 minutes were excluded to ensure the quality of questionnaires (Liu et al., 2020). Ethics approval for this study was received from the Ethics Committee of the Naval Medical University. Informed consent was obtained from all participants.

### Procedure

In view of the situation of the outbreak and the purpose of this study, we designed and published an anonymous questionnaire survey on Sojump (www.sojump.com), a network platform similar to Mechanical Turk that is operated by Amazon. After the purpose of the survey was explained, participants were ensured that all answers and identifying information would be kept confidential. Participants who volunteered to fill out the survey and hand in would receive a brief feedback about their personal mental health condition based on their answers. Afterwards, all data would be automatically summarized offline for further analysis.

### Measures

A questionnaire about socio-demographic information was designed to collect personal information (gender, age, education background), current location, recent epidemic area contact history (having a history of residence in or travel to Wuhan, or contacting with someone returning from Wuhan during the last 14 days), the classification of population (the general public, health care workers, close contacts, confirmed or suspected patients) which was referred to the risk of infection, and subjective sleep quality. For subjective sleep quality, participants were asked to rate their sleep quality during the past month on a rating scale ranging from 0 (very good) to 3 (very bad). Single-item sleep quality assessment has been identified as valid in previous study (Cappelleri et al., 2009).

PTSD symptoms were assessed by a 20-item self-report measure, the PTSD Checklist for DSM-5 (PCL-5) (Weathers et al., 2013b), which was used here to evaluate the degree of people being impacted by COVID-19 over the past month. Each item of the measure was rated from 0 (not at all) to 4 (extremely), representing the degree to which an individual has been bothered by trauma-related symptoms. A total score was obtained by summing the 20 items, ranging from 0 to 80, and higher scores indicated higher level of PTSD symptoms. In this study, a PCL-5 score of 33 or higher was regarded as a possible PTSD case (Bovin et al., 2016). Meanwhile, the 20 items of PCL-5 could be classified into four symptom clusters corresponding to Diagnostic and Statistical Manual of Mental Disorders, 5th edition (DSM-5): intrusive experience (Criterion B, item 1-5), persistent avoidance (Criterion C, item 6-7), negative alterations in cognitions and mood (Criterion D, item 8-14), and marked alterations in arousal and reactivity (Criterion E, item 15-20). Each item rated as 2 (moderately) or higher was regarded as a symptom endorsed, and symptomatic assessment criteria for a probable PTSD was at least 1 B, 1 C, 2 D, and 2 E. The Cronbach’s α of PCL-5 was 0.91 in the current sample.

### Statistical Analysis

Statistical analysis was performed using SPSS 21.0 for Windows. A two-tailed test was used and the significance level was set at *p* < 0.05. The scores of PCL-5 were expressed as mean ± SD. All possible associated factors were expressed as categorical variables. First, descriptive statistics for socio-demographic variables was conducted. The percentage of participants who met probable PTSD diagnosis was also examined. Then, t test and one-way analysis of variance were performed to test whether differences in socio-demographic characteristics accounted for the severity of PTSD. Finally, multiple linear regression analysis (enter) was conducted to identify the risk factors for PTSD, with the score of PCL-5 entered as the dependent variable and potential associated variables entered as independent variables.

## Results

The process of enrollment of study participants was displayed in **Figure 1**. Given that there were 7711 confirmed cases of COVID-19 in the mainland of China by 29 January 2020, the estimated minimum sample size was 1536 with an alpha of 0.05 and a power of 90%. Totally, the number of returned questionnaires was 2126, and 35 were excluded due to response time < 2 minutes or > 30 minutes. Finally, 2091 participants were recruited in this survey, with a valid response rate of 98.4%.

**Figure 1.**
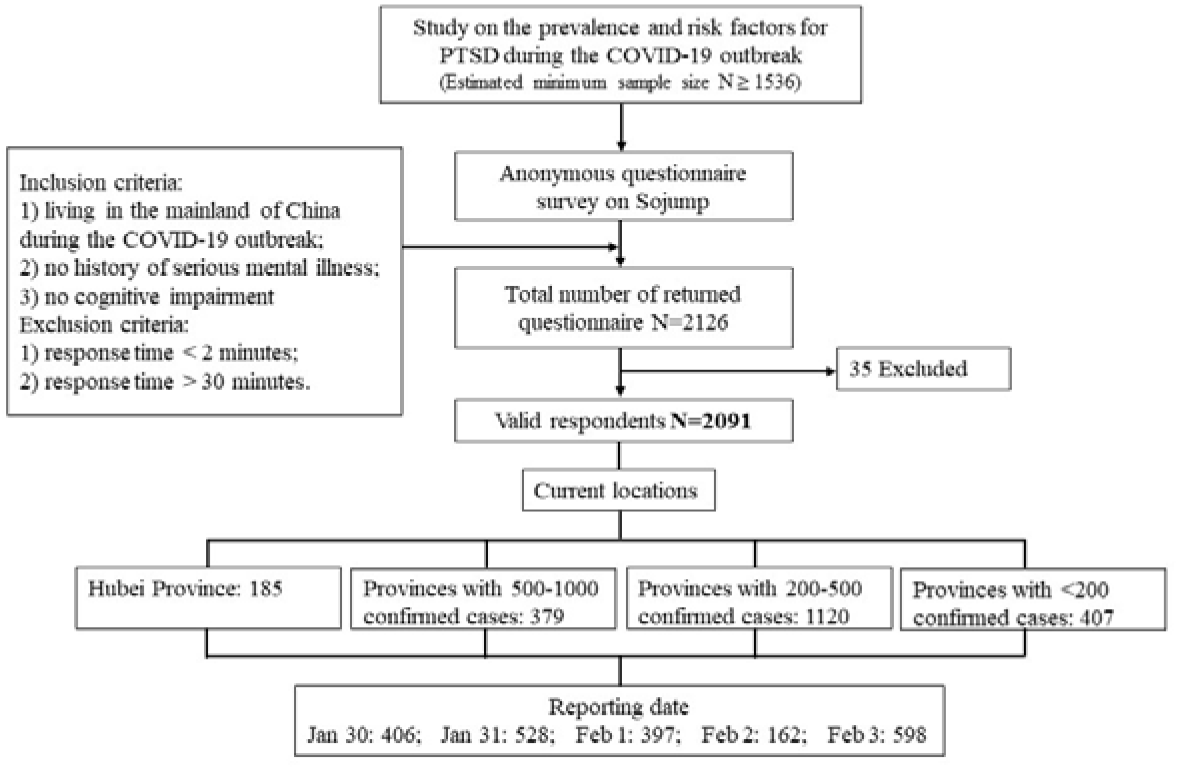
Flow chart of the enrollment of study participants.

### Sample socio-demographic characteristics

The socio-demographic characteristics are presented in **Table 1**. Among the 2091 participants, 1272 (60.8%) were female. In consideration of large age span, the participants were divided into 6 different age groups in an interval of 10 years: 30 (1.4%) were under 18, 659 (31.5%) were between 18 and 29, 615 (29.4%) were between 30 and 39, 555 (26.5%) were between 40 to 49, 180 (8.6%) were between 50 to 59, and 52 (2.5%) were over 60. Of the participants, 261 (12.5%) received an education of high school or below, 1351 (64.6%) were university or college, and 479 (22.9%) were postgraduate or above. The participants’ current locations were sorted into four groups by the number of confirmed cases of each province (by 3 February): Hubei Province (8.8%), provinces with 500-1000 confirmed cases (18.1%), provinces with 200-500 confirmed cases (53.6%), and provinces with confirmed cases less than 200 (19.5%). The date of participants filling out the survey was recorded, and there were 406 (19.4%), 528 (25.3%), 397 (19.0%), 162 (7.7%), and 598 (28.6%) participants from 30 January to 3 February, respectively. Among all participants, 240 (11.5%) had recent epidemic area contact history. In terms of the classification of population, the majority were the general public (82.4%) followed by health care workers (15.3%), while only 2.3% were suspected or confirmed patients and people having close contact with patients. Based on the risk of infection, the participants were reclassified into three groups: low-risk public (the general public), high-risk public (suspected or confirmed patients and people having close contact with patients), and health care workers. As for subjective sleep quality, 842 (40.3%) reported very good, 890 (42.6%) reported good, 326 (15.6%) reported bad, and 33 (1.6%) reported very bad.

**Table 1.** Socio-demographic characteristics of the participants

|  | N | % |
| --- | --- | --- |
| **Age** |  |  |
| <18 | 30 | 1.4 |
| 18-29 | 659 | 31.5 |
| 30-39 | 615 | 29.4 |
| 40-49 | 555 | 26.5 |
| 50-59 | 180 | 8.6 |
| ≥60 | 52 | 2.5 |
| **Gender** |  |  |
| Male | 819 | 39.2 |
| Female | 1272 | 60.8 |
| **Education background** |  |  |
| High school or below | 261 | 12.5 |
| University or college | 1351 | 64.6 |
| Postgraduate or above | 479 | 22.9 |
| **Current location** |  |  |
| Hubei | 185 | 8.8 |
| Provinces with 500-1000 confirmed cases | 379 | 18.1 |
| Provinces with 200-500 confirmed cases | 1120 | 53.6 |
| Provinces with < 200 confirmed cases | 407 | 19.5 |
| **Reporting date** |  |  |
| Jan 30,2020 | 406 | 19.4 |
| Jan 31,2020 | 528 | 25.3 |
| Feb 1,2020 | 397 | 19.0 |
| Feb 2,2020 | 162 | 7.7 |
| Feb 3,2020 | 598 | 28.6 |
| **Epidemic area contact history** |  |  |
| No | 1851 | 88.5 |
| Yes | 240 | 11.5 |
| **Classification of population** |  |  |
| Health care workers | 320 | 15.3 |
| Low-risk public | 1722 | 82.4 |
| High-risk public | 49 | 2.3 |
| **Subjective sleep quality** |  |  |
| Very good | 842 | 40.3 |
| Good | 890 | 42.6 |
| Bad | 326 | 15.6 |
| Very bad | 33 | 1.6 |
| **Total** | 2091 | 100.0 |

### Prevalence of PTSD

Among all participants enrolled in this study, 96 reported high level of PTSD symptoms, with a PCL-5 score of 33 or higher, indicating that the prevalence of PTSD in this sample was 4.6%. As a rough estimate, the prevalence of PTSD in low-risk public was 5.2%, in high-risk public was 18.4%, and in health care workers was 4.4%. Besides, the prevalence of PTSD was 5.3% by symptomatic assessment criteria, which was slightly higher than the prevalence estimated by the score threshold of 33. As for participants meeting both the threshold and symptomatic assessment criteria, the prevalence of PTSD was 3.3%. The total score of PCL-5 was used for further analysis.

### Associated factors for the severity of PTSD

**Table 2** presents the group differences of socio-demographic factors in the PCL-5 scores. There were no significant difference between the scores of participants with different education background (F = 1.329, *p* = 0.265), or that of all ages (F = 1.912, *p* = 0.089). Statistically significant differences were observed between different gender (t = −5.227, *p* < 0.001), and female reported higher PTSD symptoms. The participants in Hubei province also showed statistically differences in their PCL-5 scores (F = 13.263, *p* < 0.001). It should be noted that the different date when participants filled out the questionnaire also showed significant differences in PCL-5 scores (F = 4.018, *p* = 0.003). In the aspect of subjective sleep quality, participants with worse sleep quality scored higher on PCL-5 (F = 185.707, *p* < 0.001). With regard to the epidemic area contact history and classification of population, both of which were connected with the risk of infection, it was demonstrated significant differences between the scores of participants who had epidemic area contact history or not (*p* < 0.01), as well as participants belonging to different classifications of population (*p* < 0.001). Furthermore, as is illustrated in **Figure 2**, participants who had epidemic area contact history reported higher PCL-5 scores, and high-risk public had the highest PCL-5 scores in comparison with low-risk public and health care workers. Overall, gender, current location, reporting date, subjective sleep quality, epidemic area contact history, and classification of population could be considered as associated factors for the severity of PTSD.

### Predictive and risk factors for PTSD

Multiple linear regression analysis revealed that participant’s age and education background were not related to their PTSD symptoms during the outbreak. Nevertheless, it was noteworthy that gender (*p* < 0.001), epidemic area contact history (*p* = 0.047), classification of population (*p* < 0.001), and subjective sleep quality (*p* < 0.001) could be regarded as predictive factors for PTSD. Females and those having epidemic area contact history reported higher level of PTSD. High-risk public were most likely to develop PTSD, followed by low-risk public and health care workers. As for subjective sleep quality, the worse the sleep quality, the higher level the PTSD symptoms. **Table 3** shows the results of the regression analysis. In summary, the amount of total variation accounted for by these variables in the PCL-5 scores was significant (R^2^ = 23.2%, F = 180.835, *p* < 0.001).

**Table 2.** Group differences in the scores on the PCL-5

|  | PCL-5 Scores |  | *F/t* | *p*-value |
| --- | --- | --- | --- | --- |
|  | Mean | SD |  |  |
| **Age** |  |  | 1.912 | 0.089 |
| <18 | 15.13 | 15.62 |  |  |
| 18-29 | 11.18 | 9.67 |  |  |
| 30-39 | 12.37 | 10.83 |  |  |
| 40-49 | 11.59 | 10.02 |  |  |
| 50-59 | 12.03 | 10.38 |  |  |
| ≥60 | 9.90 | 8.31 |  |  |
| **Gender** |  |  | -5.227 | <0.001 |
| Male | 10.33 | 9.26 |  |  |
| Female | 12.64 | 10.77 |  |  |
| **Education background** |  |  | 1.329 | 0.265 |
| High school or below | 11.64 | 10.41 |  |  |
| University or college | 11.98 | 10.57 |  |  |
| Postgraduate or above | 11.10 | 9.25 |  |  |
| **Current location** |  |  | 13.263 | <0.001 |
| Hubei | 15.95 | 12.73 |  |  |
| Provinces with 500-1000 confirmed cases | 11.09 | 9.03 |  |  |
| Provinces with 200-500 confirmed cases | 11.73 | 10.50 |  |  |
| Provinces with < 200 confirmed cases | 10.44 | 8.87 |  |  |
| **Reporting date** |  |  | 4.018 | 0.003 |
| Jan 30,2020 | 11.05 | 10.42 |  |  |
| Jan 31,2020 | 10.56 | 8.86 |  |  |
| Feb 1,2020 | 12.35 | 10.78 |  |  |
| Feb 2,2020 | 13.06 | 11.87 |  |  |
| Feb 3,2020 | 12.47 | 10.38 |  |  |
| **Subjective sleep quality** |  |  | 185.707 | <0.001 |
| Very good | 7.85 | 7.50 |  |  |
| Good | 11.82 | 8.78 |  |  |
| Bad | 19.43 | 12.20 |  |  |
| Very bad | 32.58 | 17.25 |  |  |

**Table 3.**
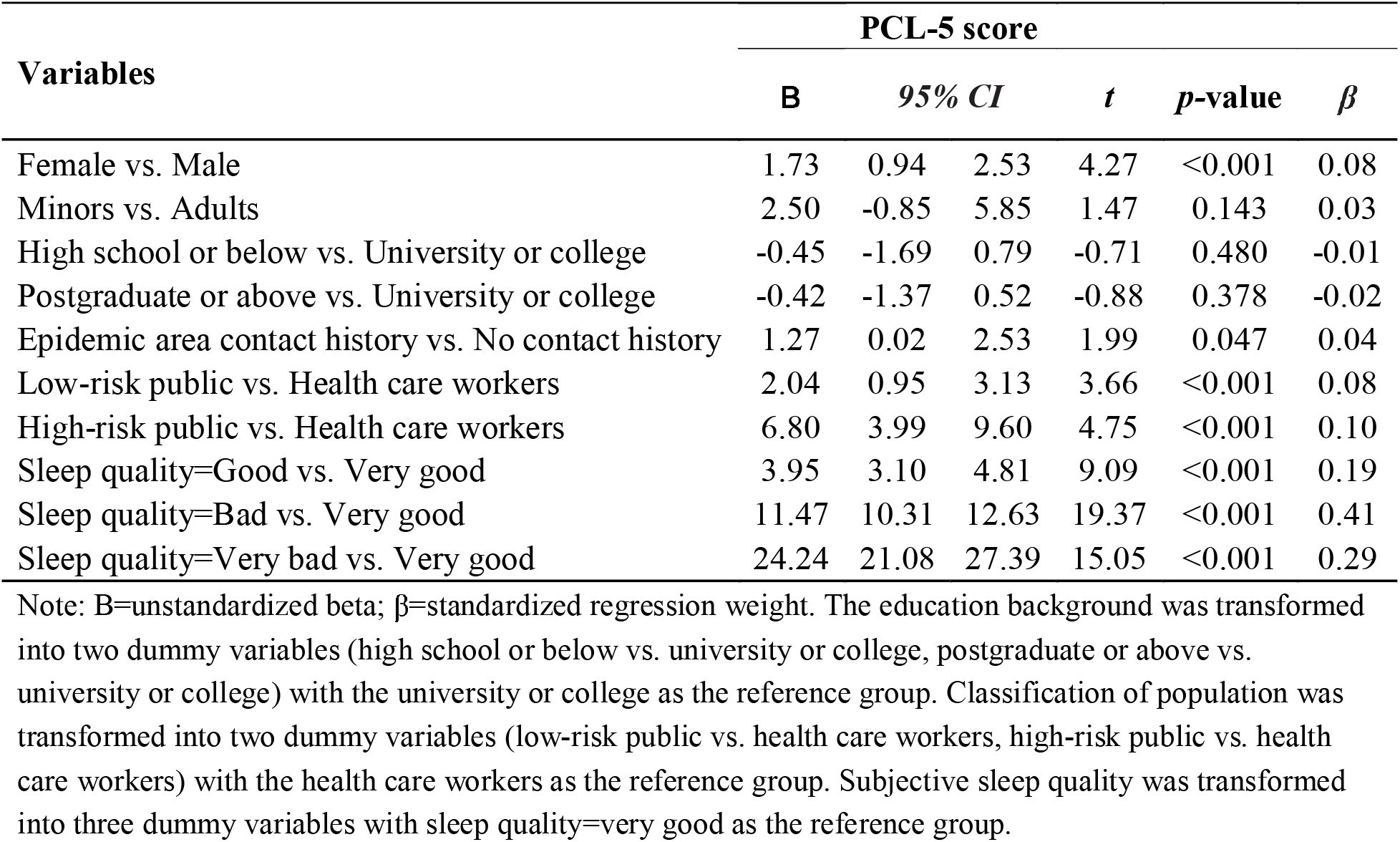
Regression Analysis with PCL-5 Score as the Dependent Variable (n = 2091)

**Figure 2.**
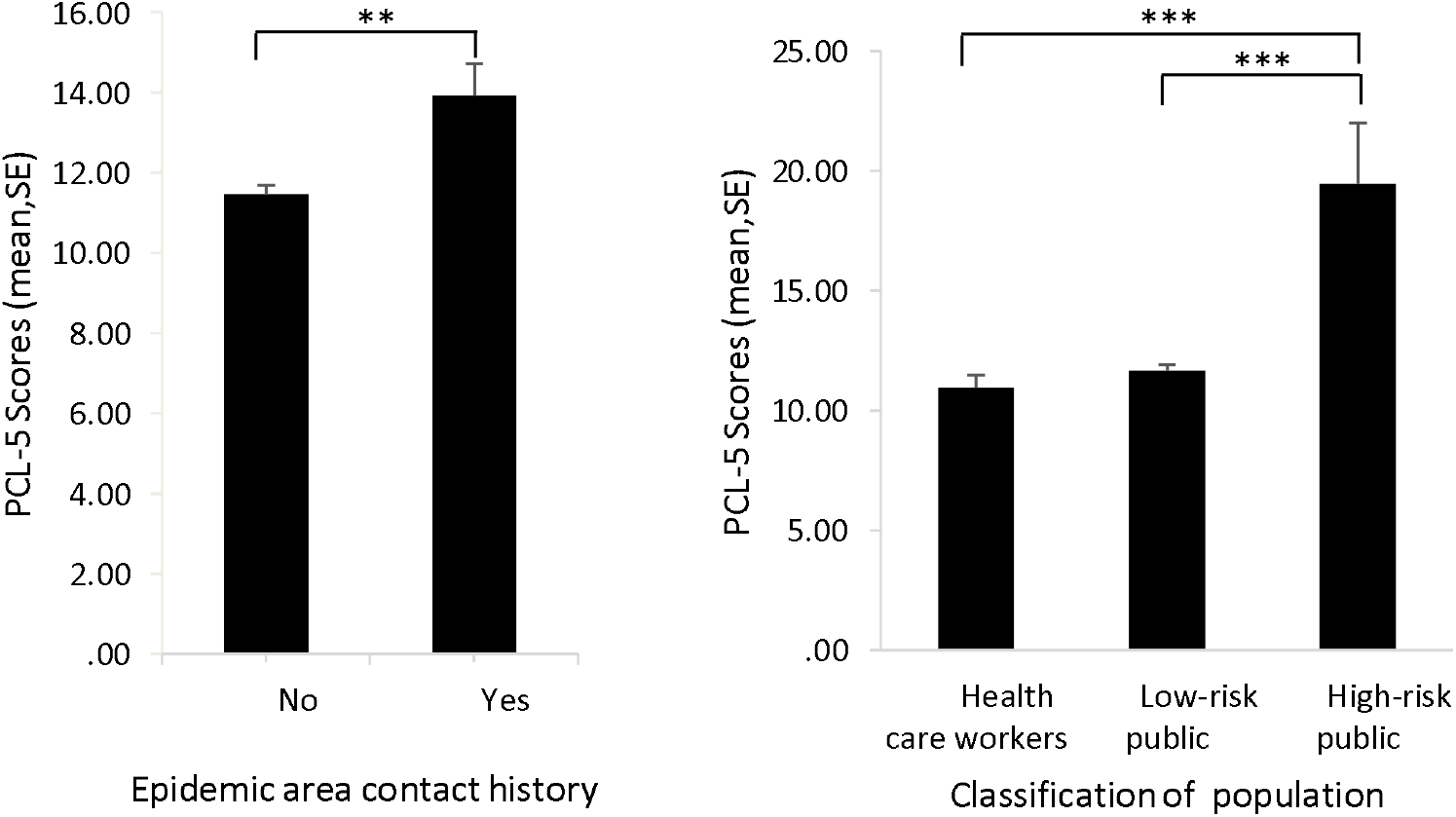
Group differences of PCL-5 scores in epidemic area contact history and classification of population. Note: ^**^, *p*-value < 0.01; ^***^, *p*-value < 0.001.

## Discussion

The increasing number of new COVID-19 cases and its rapidity of human-to-human transmission have attracted public attention, and countries around the world are concerned about the further spread of the virus. The results from this study revealed that 4.6% of the respondents might have PTSD. Gender, current location, reporting date, subjective sleep quality, epidemic area contact history, and classification of population might be associated factors for the severity of PTSD. Furthermore, female, having recent epidemic area contact history, population at high risk of infection, and poor sleep quality were identified as risk factors for PTSD.

In the current study, females were prone to developing higher levels of PTSD, which was in line with a previous study exploring the predictors for PTSD during the H1N1 epidemic (Xu et al., 2011). Moreover, as reported by some literature, females are about twice as likely to develop PTSD (Christiansen and Elklit, 2012; Kessler, 1995). With regard to the reason, it is suggested that females tend to experience higher levels of potential risk factors, like depression, physical anxiety sensitivity, and helplessness (Christiansen and Hansen, 2015). Accordingly, the female respondents in this study showed higher PTSD than the male respondents.

As expected, poor sleep quality was a significant predictive factor for PTSD, which has been well-documented in relevant studies (Elissa et al., 2019; Janna et al., 2018; Noel et al., 2017). Previous studies have shown that subjective sleep disturbances and disruption of REM sleep during the early aftermath of trauma can predict future development of PTSD (Mellman et al., 2002; Mellman et al., 2007). Moreover, sleep disturbances are a core characteristic of PTSD (Spoormaker and Montgomery, 2008). For the majority of general public during the COVID-19 outbreak, poor sleep quality tended to be recognized more easily and gain more attention than PTSD symptoms, which might assist in the identification of PTSD and shed light on the psychological interventions for people affected by the COVID-19 pandemic.

People who resided in Hubei Province, the original and worst disaster-affected area, reported the highest level of PTSD. This result was consistent with a study about the psychological impact of SARS, which demonstrated that residents in high SARS-prevalent locations were prone to developing probable PTSD (Lee et al., 2006). On the one hand, people living in high infection-prevalent locations perceived themselves to be at higher risk of infection; on the other hand, the lockdown of transport hub in most cities of Hubei Provinces led to negative psychological impact on residents in those cities.

Notably, the epidemic area contact history determined the risk of infection to some extent in the present study, but there is no consensus regarding this relation. Alexander and Klein suggested that those people who were repeatedly exposed to traumatic events were prone to suffering all kinds of psychological problems (Alexander and Klein, 2001). On the contrary, Declercq et al. found no relationship between the frequency of encountering critical incidents and the occurrence of PTSD symptoms (Declercq et al., 2011). Jiahong et al. indicated that knowledge on H1N1 and exposure to H1N1 patients could not predict stress symptoms (Xu et al., 2011). Further research is needed to explore the influence of epidemic exposure history on the development of PTSD.

In terms of the classification of population, high-risk public, including confirmed or suspected patients and those who had close contacts with patients, showed the most severe PTSD symptoms. It should also be noted that the prevalence of PTSD among the confirmed or suspected patients experiencing the virus in this study was 25%. It can be inferred that their psychological condition was susceptible to being negatively impacted by the pandemic with the fear and uncertainty of the new emerging infection and without effective vaccines or medical treatments. Besides, both high-risk or low-risk public were inevitably under quarantine during that period, which might further impact their psychological consequences. A study found that quarantined people developed various distress symptoms, and the distress was positively correlated to the duration of quarantine (Hawryluck et al., 2004). Compared with the estimated rate of morbidity in health workers at the initial phase of SARS outbreak, which was 75.3% (Chong et al., 2004), and the percentage of medical staff who reported high levels of PTSD symptoms during SARS in another study, which was 10% (Wu et al., 2009), the prevalence of PTSD in health care workers participating in the present study was lower. There may be two reasons accounting for this. First, China had experience two waves of major infectious disease since 20^th^ century, as a result of which considerable experience on coping with the epidemic has been gathered. Second, health care workers generally know more about the virus than the general public, and thus they were deployed in an orderly way and commenced to work promptly with more confidence and courage to defeat the virus.

The results also revealed that the levels of PTSD reported on different date were significantly different, and participants’ PTSD symptoms seemingly got worse during the survey period. As has been mentioned above, the COVID-19 pandemic continuously escalated in China from the end of January 2020 to the beginning of February 2020. Although the trend was not obvious enough due to the short time span, it could be proved with caution that the severity of PTSD was largely influenced by the scale of the outbreak.

Inconsistent with the hypothesis, age and education background were found to be not connected with PTSD. One possible reason might be that the majority of participants enrolled in this study were not representative because they were between 18 and 60 years old with a degree from university or college. In addition, most Chinese were under home quarantine as soon as the infection worsened, so they received almost the same information regarding the epidemic, which might reduce the impact of age and education background on their PTSD symptoms.

### Clinical implications

The current study found that the public in the mainland of China showed PTSD symptoms one month after the COVID-19 outbreak, and identified several risk factors for PTSD including female, having recent epidemic area contact history, population at high risk of infection, and poor sleep quality. Thus, early psychological intervention should be implemented for the public during the COVID-19 pandemic and future public health emergencies. Most importantly, the predictors identified in this study may be useful in defining high-risk group in future epidemics and may contribute to providing early-stage psychological intervention.

### Limitations

The results of this study should be considered with several potential limitations. First, the majority of participants in this study were the general public, with confirmed or suspected patients being a small part, whose PTSD symptoms might be far more severe. Thus, more attention should be paid to the psychological condition of COVID-19 patients. Second, the measurement of PTSD in this study might be vulnerable to selection bias because of an online self-report study, such as participants’ recruitment. The potential bias might result from the fact that the elderly and people who reside in remote areas have less access to the Internet. Additionally, some participants might be reluctant to report their real psychological status in a questionnaire. Third, the prevalence of PTSD in this study was estimated by an online questionnaire rather than a clinical interview. Although online survey was the best choice during that period when a series of restrictive measures (e.g. lockdowns, stay-at-home order) limited data collection, future study can benefit from adopting a more rigorous approach with more valid and reliable measures than online surveys.

## Conclusion

Despite these limitations, several important conclusions emerge from this study. To summarize, this study firstly found that the prevalence of PTSD among the public in the mainland of China one month after the COVID-19 outbreak was 4.6%. Moreover, female, having recent epidemic area contact history, population at high risk of infection, and poor sleep quality were identified as risk factors for PTSD. When implementing psychological interventions for the public during the pandemic, particular importance should be attached to females and those at high risk of infection, such as people residing in high disease-prevalent regions or having had close contact with COVID-19 patients.

## Data Availability

The data used to support the findings of this study are available from the corresponding author upon request.

## Author contributions

LS, ZS, LW, ZZ, contributing to the writing and statistical analyses of this article, are listed as co-first authors. WL, the corresponding author, led the whole study, including putting forward this study and carrying out the study. FZ, ZS, YJ, JG, YZ, YW and NL contributed to performing the investigation and collecting all data.

We are all accountable for all aspects of the work in ensuring that questions related to the accuracy or integrity of any part of the work are appropriately investigated and resolved.

## Acknowledgements

The authors would like to acknowledge the volunteers who participated in the study and Xiaoqian Yu Ph.D. from University of South Florida, USA, for language editing.

## References

Alexander, D.A., Klein, S., 2001. Ambulance personnel and critical incidents - Impact of accident and emergency work on mental health and emotional well-being. Br J Psychiatry 178, 76–81.

Alexander, E.G., Susan, C.B., Ralph, S.B., Raoul, J.d.G., Christian, D., Anastasia, A.G., Bart, L.H., Chris, L., Andrey, M.L., Benjamin, W.N., Dmitry, P., Stanley, P., Leo, L.M.P., Dmitry, S., Igor, A.S., Isabel, S., John, Z., 2020. Severe acute respiratory syndrome-related coronavirus – The species and its viruses, a statement of the Coronavirus Study Group.. bioRxiv.

Batterham, J. P., 2014. Recruitment of mental health survey participants using Internet advertising: content, characteristics and cost effectiveness. International Journal of Methods in Psychiatric Research 23, 184–191.

Bovin, M.J., Marx, B.P., Weathers, F.W., Gallagher, M.W., Rodriguez, P., Schnurr, P.P., Keane, T.M., 2016. Psychometric properties of the PTSD Checklist for Diagnostic and Statistical Manual of Mental Disorders-Fifth Edition (PCL-5) in veterans. Psychol Assess 28, 1379–1391.

Cappelleri, J.C., Bushmakin, A.G., McDermott, A.M., Sadosky, A.B., Petrie, C.D., Martin, S., 2009. Psychometric properties of a single-item scale to assess sleep quality among individuals with fibromyalgia. Health Qual Life Outcomes 7, 54.

Chong, M.-Y., Wang, W.-C., Hsieh, W.-C., Lee, C.-Y., Chiu, N.-M., Yeh, W.-C., Huang, T.-L., Wen, J.-K., Chen, C.-L., 2004. Psychological impact of severe acute respiratory syndrome on health workers in a tertiary hospital. British Journal of Psychiatry 185, 127–133.

Christiansen, D.M., Elklit, A., 2012. Sex differences in PTSD. In A. Lazinica & E. Ovuga (Eds.), Posttraumatic stress disorder in a global context (pp. 113-142). Rijeka, Croatia: InTech Open Access Book.

Christiansen, D.M., Hansen, M., 2015. Accounting for sex differences in PTSD: A multi-variable mediation model. Eur J Psychotraumatol 6, 26068.

Declercq, F., Meganck, R., Deheegher, J., Hoorde, H., 2011. Frequency of and subjective response to critical incidents in the prediction of PTSD in emergency personnel. J Trauma Stress 24, 133–136.

Elissa, McCarthy, Jason, DeViva, Sonya, Norman Steven, 2019. Self-assessed sleep quality partially mediates the relationship between PTSD symptoms and functioning and quality of life in U.S. Veterans: Results from the National Health and Resilience in Veterans Study. Psychological trauma: theory, research, practice and policy.

Forte, G., Favieri, F., Tambelli, R., Casagrande, M., 2020. COVID-19 Pandemic in the Italian Population: Validation of a Post-Traumatic Stress Disorder Questionnaire and Prevalence of PTSD Symptomatology. Int J Environ Res Public Health 17.

Hawryluck, L., Gold, W.L., Robinson, S., Pogorski, S., Styra, R., 2004. SARS Control and Psychological Effects of Quarantine, Toronto, Canada. Emerging Infectious Diseases 10, 1206–1212.

Janna, M., Helms, S.M., Weymann, K.B., Capaldi, V.F., Lim, M.M., 2018. Sleep Quality and Emotion Regulation Interact to Predict Anxiety in Veterans with PTSD. Behavioural Neurology 2018, 1–10.

Juraschek, S.P., Plante, T.B., Charleston, J., Miller, E.R., Hermosilla, M., 2018. Use of online recruitment strategies in a randomized trial of cancer survivors. Clinical Trials 15, 174077451774582.

Kessler, R.C., 1995. Posttraumatic Stress Disorder in the National Comorbidity Survey. Arch.gen.psychiatry 52, 1048.

Kessler, R.C., 2000. Posttraumatic stress disorder: The burden to the individual and to society. Journal of Clinical Psychiatry 61 Suppl 5, 4-12; discussion 13-14.

Leach, L.S., Butterworth, P., Poyser, C., Batterham, P.J., Farrer, L.M., 2017. Online Recruitment: Feasibility, Cost, and Representativeness in a Study of Postpartum Women. 19, e61.

Lee, T.M., Chi, I., Chung, L.W., Chou, K.L., 2006. Ageing and psychological response during the post-SARS period. Aging Ment Health 10, 303–311.

Lin, C.Y., Peng, Y.C., Wu, Y.H., Chang, J., Chan, C.H., Yang, D.Y., 2007. The psychological effect of severe acute respiratory syndrome on emergency department staff. Emerg Med J 24, 12–17.

Liu, N., Zhang, F., Wei, C., Jia, Y., Shang, Z., Sun, L., Sun, Z., Zhou, Y., Wang, Y., Liu, W., 2020. Prevalence and predictors of PTSS during the COVID-19 Outbreak in China Hardest-hit Areas: Gender differences matter. Psychiatry Research 112921.

Mak, I.W.C., Chu, C.M., Pan, P.C., Yiu, M.G., Chan, V.L., 2009. Long-term psychiatric morbidities among SARS survivors. 31, 318–326.

Mellman, T.A., Bustamante, V., Fins, A.I., Pigeon, W.R., Nolan, B., 2002. REM sleep and the early development of posttraumatic stress disorder. Am J Psychiatry 159, 1696–1701.

Mellman, T.A., Pigeon, W.R., Nowell, P.D., Nolan, B., 2007. Relationships between REM sleep findings and PTSD symptoms during the early aftermath of trauma. J Trauma Stress 20, 893–901.

Munster, V., Koopmans, M., van Doremalen, N., van Riel, D., de Wit, E., 2020. A novel coronavirus emerging in China — key questions for impact assessment.. N Engl J Med.

Noel, M., Vinall, J., Tomfohr-Madsen, L., Holley, A.L., Wilson, A.C., Palermo, T.M., 2017. Sleep Mediates the Association between PTSD Symptoms and Chronic Pain in Youth. Journal of Pain, S1526590017307186.

Quach, S., Pereira, J.A., Russell, M.L., Wormsbecker, A.E., Kwong, J., 2013. The Good, Bad, and Ugly of Online Recruitment of Parents for Health-Related Focus Groups: Lessons Learned. Journal of Medical Internet Research 15, e250.

Spoormaker, V.I., Montgomery, P., 2008. Disturbed sleep in post-traumatic stress disorder: Secondary symptom or core feature? Sleep Medicine Reviews 12, 169–184.

The 2019-nCoV Outbreak Joint Field Epidemiology Investigation Team, Li, Q., 2020. Notes from the field: an outbreak of NCIP (2019-nCoV) infection in China — Wuhan, Hubei Province, 2019–2020. China CDC Weekly 2, 79–80.

Wang, C., Horby, P.W., Hayden, F.G., Gao, G.F., 2020. A novel coronavirus outbreak of global health concern. The Lancet.

Weathers, F.W., Litz, B.T., Keane, T.M., Palmieri, P.A., Marx, B.P., Schnurr, P.P., 2013a. The PTSD Checklist for DSM-5 (PCL-5) – LEC-5 and Extended Criterion A [Measurement instrument]. Available from https://www.ptsd.va.gov/.

Weathers, F.W., Litz, B.T., Keane, T.M., Palmieri, P.A., Marx, B.P., Schnurr, P.P., 2013b. The PTSD Checklist for DSM-5 (PCL-5). Scale available from the National Center for PTSD at www.ptsd.va.gov.

WHO, 2020. WHO Director-General’s remarks at the media briefing on 2019-nCoV on 11 February 2020. Geneva: World Health Organization, 2020. (https://www.who.int/dg/speeches/detail/who-director-general-s-remarks-at-the-media-briefing-on-2019-ncov-on-11-february-2020) (assessed 12 Feb, 2020).

WHO, 2021. Coronavirus (COVID-19), 17 January 2021. Geneva: World Health Organization, 2021 (https://covid19.who.int/) (assessed 18 Jan, 2021).

Wu, K.K., Chan, S.K., Ma, T.M., 2005. Posttraumatic stress, anxiety, and depression in survivors of severe acute respiratory syndrome (SARS). J Trauma Stress 18, 39–42.

Wu, P., Fang, Y., Guan, Z., Fan, B., Kong, J., Yao, Z., Liu, X., Fuller, C.J., Susser, E., Lu, J., 2009. The psychological impact of the SARS epidemic on hospital employees in China: exposure, risk perception, and altruistic acceptance of risk. 54, 302.

Xu, J., Zheng, Y., Wang, M., Zhao, J., Cheng, Y., 2011. Predictors of symptoms of posttraumatic stress in Chinese university students during the 2009 H1N1 influenza pandemic. Medical Science Monitor International Medical Journal of Experimental & Clinical Research 17, PH60–64.

Zhu, N., Zhang, D., Wang, W., Li, X., Yang, B., Song, J., Zhao, X., Huang, B., Shi, W., Lu, R., Niu, P., Zhan, F., Ma, X., Wang, D., Xu, W., Wu, G., Gao, G.F., Tan, W., China Novel Coronavirus, I., Research, T., 2020. A Novel Coronavirus from Patients with Pneumonia in China, 2019. N Engl J Med.

